# Tumor-specific activity of precision medicines in the NCI-MATCH trial

**DOI:** 10.1101/2023.03.30.23287951

**Authors:** Ivvone Zhou, Deborah Plana, Adam C. Palmer

## Abstract

**Background:** NCI-MATCH is a precision medicine basket trial designed to test the effectiveness of treating cancers based on specific genetic changes in patients’ tumors, regardless of cancer type. Multiple subprotocols have each tested different targeted therapies matched to specific genetic aberrations. Most subprotocols exhibited low rates of tumor shrinkage as evaluated across all tumor types enrolled. We hypothesized that these results may arise because these precision cancer therapies have tumor type-specific efficacy, as is common among other cancer therapies.

**Methods:** To test the hypothesis that certain tumor types are more sensitive to specific therapies than other tumor types, we applied permutation testing to tumor volume change and progression-free survival data from ten published NCI-MATCH subprotocols (together n=435 patients). False discovery rate was controlled by the Benjamini-Hochberg procedure.

**Results:** Six of ten subprotocols exhibited statistically significant evidence of tumor-specific drug sensitivity, four of which were previously considered negative based on response rate across all tumors. This signal-finding analysis highlights potential uses of FGFR tyrosine kinase inhibition in urothelial carcinomas with actionable *FGFR* aberrations, MEK inhibition in lung cancers with *BRAF* non-*V600E* mutations, and MEK inhibition in cholangiocarcinomas with *NRAS* mutations.

**Conclusions:** These findings support the value of basket trials because even when precision medicines do not have tumor-agnostic activity, basket trials can identify tumor-specific activity for future study.

## Introduction

Oncogenes drive the growth of many cancers, making their products a popular target for drug development^1^. The success of drugs like imatinib, which inhibits the BCR-ABL fusion protein in chronic myeloid leukemia (CML)^2^, has spurred the rise of molecularly-targeted therapies and the paradigm of using tumor genetics to identify patients who will respond to those therapies. Under this paradigm, it is possible that tumors of different tissue types but the same molecular aberrations may be responsive to the same drug^3^. This approach has produced a need for specialized clinical trial designs to evaluate the effectiveness of drugs across multiple tumor types^4^.

Basket trials enroll multiple subgroups of patients to study the effect of a specific treatment on multiple types of cancer^5^. Commonly, patients are enrolled on the basis of a tumor biomarker that is hypothesized to confer sensitivity to the treatment (e.g. *NTRK* fusion for NTRK inhibition), and enrollment can include cancers from a variety of tissues of origin (e.g. lung, colon, etc). Basket trials can be designed to operate as multiple, independent tissue-specific trials^6^, or, they may evaluate efficacy in the overall trial population^7,8^, which has recently led to ‘tumor-agnostic’ drug approvals. Notable examples include the approval of larotrectinib for *NTRK*-fusion tumors, and pembrolizumab for microsatellite instability-high or mismatch repair deficient tumors^9,10^.

The National Cancer Institute Molecular Analysis for Therapy Choice (NCI-MATCH) is a basket trial that matches patients to targeted therapies on the basis of genetic abnormalities in tumors (e.g. mutations, fusions, amplifications)^11,12^. In this trial, patients with refractory malignancies are screened for qualifying genetic alterations in tumors, and if eligible, receive a treatment targeting that alteration as part of a Phase 2 subprotocol. Patients with a variety of cancer types are enrolled as separate “baskets” in each subprotocol testing a different drug.

The primary endpoint of each subprotocol is overall response rate (ORR), evaluated in all patients regardless of tumor type. By the cutoff date of our analysis (August 2022), 12 subprotocols have reported efficacy findings for different drugs (**Supplementary Table 1**). Organizationally this trial has been impactful by establishing the feasibility of a large multi-histology trial and treatment assignment on the basis of tumor genetics. However, most subprotocols reported thus far did not meet their primary efficacy endpoint (a response rate of at least 16%), which has been reasonably considered a disappointing result for precision oncology^13^. Subprotocol H, which evaluated dabrafenib plus trametinib in *BRAFV600E*-mutant cancers^14^, was successful enough across all tumor types to support expansion and ultimately a tumor-agnostic regulatory approval^15^. This result both confirms the value of searching for tumor-agnostic efficacy, and also shows it to be uncommon.

Viewed in historical context, the NCI-MATCH results are not unexpected, because most approved cancer therapies are effective in certain tumor types, and would exhibit low response rates if they were tested in a tumor-agnostic set of patients. Current practices for selection of cancer therapies depend heavily on tumor histology and tissue of origin, including most current precision therapies, such as EGFR inhibitors^16^ and HER2 antibodies^17^. Thus, although some genomically-guided therapies have tumor-agnostic efficacy, most appear to be beneficial in specific tumor types. This tendency for tumor-specific efficacy of cancer therapies poses a challenge in identifying the best settings for the clinical development of new genomically-guided therapies. Basket trials such as NCI-MATCH are uniquely positioned to address this challenge^4^.

Although the NCI-MATCH trial was not explicitly designed for the purpose of identifying differences among tumor types in drug sensitivity, we have recently published a method that enables such investigations^18^. In our prior work, we showed that permutation testing can compare responses across multiple tumor subtypes in basket trials, even when there are few patients per subtype. Formally, this method tests whether the results of a multi-histology trial reject the null hypothesis that there is no difference in drug sensitivity between tumor types. This method confirmed the tumor-agnostic efficacy of pembrolizumab and larotrectinib in particular genetic contexts, and identified an overlooked therapeutic opportunity for HER2 inhibition in HER2-mutant lung cancers^18^.

Here, we applied our permutation-based method to identify evidence of tumor-specific drug efficacy in the NCI-MATCH trial. We found that most subprotocols exhibited evidence that specific tumor types are significantly more sensitive to targeted therapies than other tumor types as a whole. This suggests that even when basket trials do not identify tumor-agnostic efficacy, they can identify tumor-specific uses of precision medicines.

## Methods

We analyzed Tumor Volume Change (TVC) and Progression-Free Survival (PFS) from ten subprotocols of the NCI-MATCH trial (all published up to August 2022) (**Supplementary Table 1**)^14,19–27^. Of 12 published subprotocols, two studying crizotinib had a combined accrual of nine patients and were considered unevaluable in our analysis^28^. Individual participant data on TVC and PFS was digitized from published graphs (‘waterfall plots’ and ‘swimmers plots’) as described^18^. Subgroups were defined by tumor types reported in the original publications, with exceptions noted in Figure 1.

**Figure 1:**
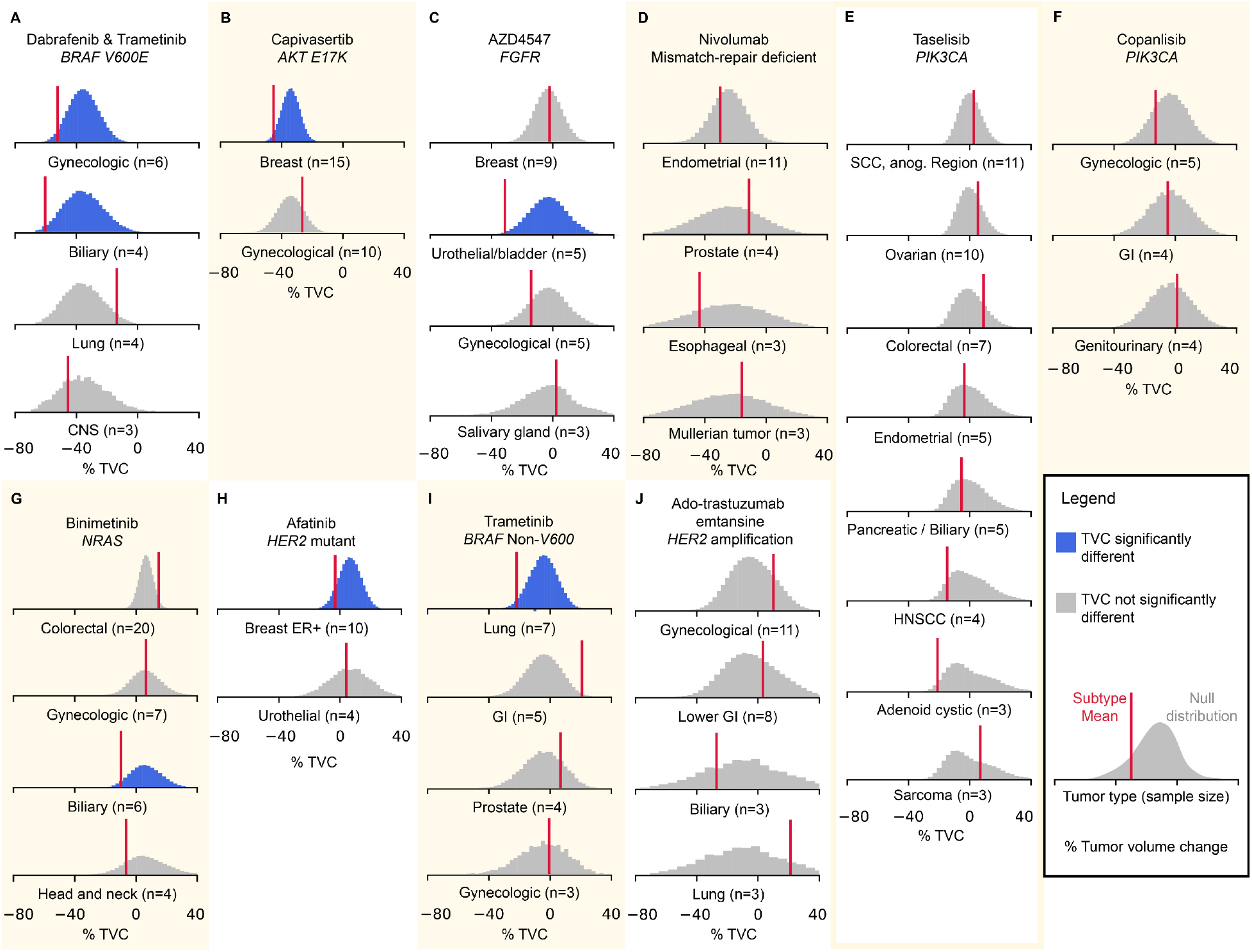
Tumor types with significant differences in tumor volume change in NCI-MATCH subprotocols. Each panel shows a distribution of tumor volume changes (TVC) under the null hypothesis of no difference between tumor types, and observed average TVC for each tumor type (red line). **A)** Dabrafenib with trametinib in *BRAFV600E*-mutated tumors. ‘Biliary’ denotes intrahepatic cholangiocarcinoma^14^. **B)** Capivasertib in *AKT E17K*-mutated tumors. We classified tumor types as gynecological, breast, or other^23^. **C**) AZD4547 in *FGFR-*mutated tumors^22^. **D)** Nivolumab in mismatch-repair deficient tumors. ‘Endometrial’ denotes Endometrial endometrioid adenocarcinoma and Endometrial endometrioid adenocarcinoma variants^20^. **E)** Taselisib in *PIK3CA*-mutated tumors. ‘SCC anog’ denotes squamous cell carcinoma of the anogenital region. **F)** Copanlisib in *PIK3CA*-mutated tumors. **G)** Binimetinib in *NRAS*-mutated tumors. **‘**Biliary’ denotes cholangiocarcinoma^24^. **H)** Afatinib in *HER2-*mutated tumors. **I)** Trametinib in *BRAF* Non-*V600E* mutated tumors^21^. **J)** Ado-trastuzumab emtansine in *HER2*-amplified tumors^19^. For all trials, subgroups of miscellaneous tumors (denoted “other”), or subgroups of fewer than three patients, were not tested for differences.

**Figure 2:**
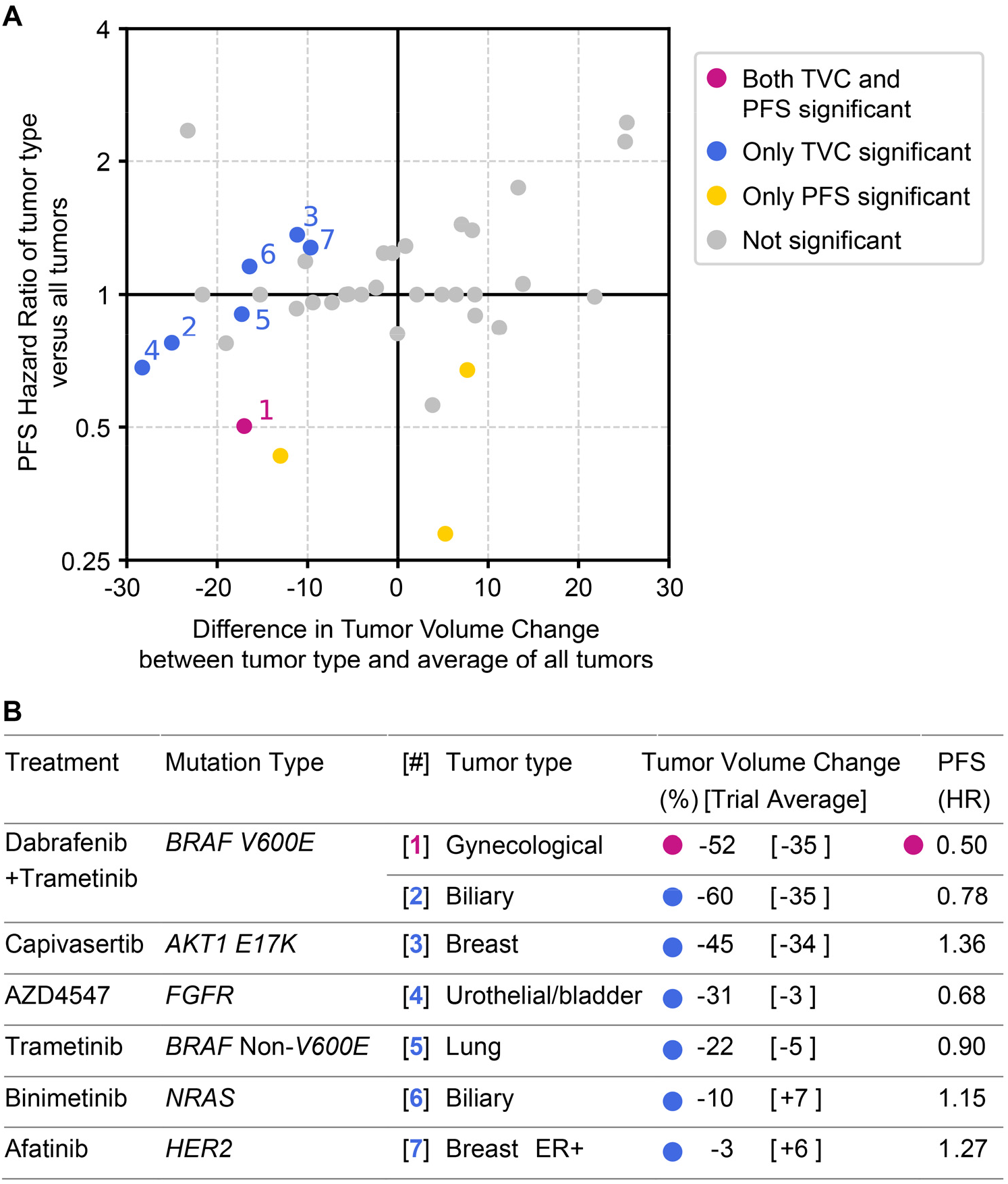
Tumor type-specific hazard ratios and differences in tumor volume change compared to each subprotocol’s average. **A)** Scatterplot of each tumor type’s Hazard Ratio (PFS of tumor type versus PFS of all tumors in a subprotocol) and difference in tumor volume change (average TVC of tumor type compared with average TVC of all tumors in a subprotocol). Note, HR of 1 means a tumor type exhibited the same PFS as that of all tumor types; it does not mean no therapeutic effect. **B)** Summary of tumor types with significantly better TVC and/or PFS compared to all tumors enrolled in a subprotocol.

We tested the null hypothesis that a given tumor type is equally sensitive to a therapy as compared to all tumor types in a trial, against the alternative hypothesis that a given tumor type is more sensitive to that therapy than other types. This hypothesis was tested by Monte Carlo permutation testing, as previously described^18^. For each tumor type’s sample size (e.g. 10 patients), that number of outcomes are randomly sampled from the pool of all outcomes, and a metric of efficacy is calculated (such as average TVC). This is repeated 10^7^ times to compose a sampling distribution of efficacy metrics under the null hypothesis that there is no difference in efficacy by tumor type. A tumor type’s empiric *P*-value for superior efficacy is the area under the null distribution to the left of that tumor type’s observed average TVC (i.e. greater tumor shrinkage). This *P*-value quantifies the probability that the null hypothesis is true (no difference in efficacy) given the observed data.

To control the false discovery rate (FDR) associated with multiple hypothesis testing, significance thresholds were defined by the Benjamini-Hochberg procedure with FDR=0.25. Multiple hypothesis correction is typically absent from multi-histology trials and FDR is not to be confused with type 1 error; simulations indicated that FDR of 25% yields a false positive (type 1 error) rate of 3%, which is more stringent than 10% false positive rate common in Simon two-stage designs^18^. As previously reported, when compared to a binomial test of response rate, permutation testing provides a superior true positive rate (power) at the same false positive rate (type 1 error)^18^.

We used TVC as the primary metric of efficacy and PFS as a secondary metric. The same permutation test with FDR correction was applied for PFS, with significance assessed for the Hazard Ratio (Cox Proportional Hazards model) of the subsample PFS events compared with all PFS events. As discussed previously^18^, PFS can be challenging to interpret in multi-histology trials because of differences in tumor growth rates. Evidence of longer PFS in a tumor type is most informative if it is concordant with greater tumor shrinkage. We opted not to draw conclusions based on PFS alone without a significant signal of greater tumor shrinkage.

## Results

Permutation testing found that six of ten subprotocols from NCI-MATCH had at least one tumor type with significantly greater tumor shrinkage than other types enrolled (**Figure 1; Supplementary Table 2**). Notably, significant evidence of tumor-specific drug efficacy was detected in four subprotocols that did not meet their primary efficacy endpoint of ≥16% objective response rate across all tumor types. Most of these signals of tumor-specific efficacy from NCI-MATCH are independently supported by other clinical trials.

Subprotocol H tested dabrafenib plus trametinib for *BRAF V600E* tumors^14^ (**Figure 1A**). With a 38% objective response rate (ORR), this was one of the most successful basket trials, resulting in a tumor-agnostic FDA approval. Even among a broadly effective treatment, permutation testing identified two tumor types that were significantly more responsive than all others. Patients with gynecological cancers (n=6; 5 being low-grade serous ovarian carcinoma; LGSOC) had greater tumor volume shrinkage (−52% versus trial average -35%; *P* = 0.036) and, at an FDR of 25% by Benjamini-Hochberg, also longer PFS versus all tumors in the trial (HR 0.5; *P* = 0.056). LGSOC is a less aggressive but chemo-resistant subtype of ovarian cancer, and therefore its longer PFS may be an innate biological feature, but the strong tumor shrinkage elicited by dabrafenib plus trametinib compares favorably to the 4% ORR of platinum chemotherapy in recurrent LGSOC^29^. Patients with intrahepatic cholangiocarcinoma (denoted biliary; n=4) also exhibited significantly greater tumor shrinkage (−60% versus trial average - 35%; *P* = 0.016). As noted by Wainberg *et al*, this signal is supported by an independent observation of 41% ORR of dabrafenib plus trametinib in *BRAF V600E* mutant biliary tract cancers^30^.

Subprotocol R tested the MEK inhibitor trametinib in solid tumors or lymphomas with *BRAF* non-*V600* mutations or fusions^21^ (**Figure 1I**). Assessed in all patients, trametinib was not effective, having an ORR of 3% and an average tumor volume change of -4%. However, patients with *BRAF* non-*V600* mutant lung cancer (n=7 evaluable for TVC) exhibited significantly greater tumor shrinkage versus all patients in the trial (−22% versus trial average - 4%; *P* = 0.026), with 3 of 7 patients (42%) experiencing greater than 30% reduction in target lesion size.

Subprotocol W tested the Fibroblast Growth Factor Receptor (FGFR) 1-3 inhibitor AZD4547 in tumors with aberrations in FGFR1-3^22^ (**Figure 1C**). AZD4547 did not meet the primary efficacy endpoint, eliciting an 8% ORR. Patients with urothelial carcinomas (n=5) had significantly greater tumor shrinkage versus all patients (−31% versus trial average -3%; *P* = 0.011). This signal of AZD4547 efficacy is consistent with the efficacy and FDA approval of the FGFR inhibitor erdafitinib for *FGFR2*-*3* altered urothelial cancers^31^.

Subprotocol Y tested the pan-AKT inhibitor capivasertib in AKT1 E17K mutant tumors and demonstrated an ORR of 29%^23^ (**Figure 1B**). The most prevalent qualifying tumor type was breast carcinomas, which demonstrated significantly greater tumor volume change versus all patients (−45% versus trial average -34%; *P* = 0.025). This signal of greater efficacy in breast carcinomas is consistent with the demonstrated efficacy and FDA approval of capivasertib combined with paclitaxel for metastatic triple-negative breast cancer, in which the largest survival improvements were observed in *PIK3CA*/*AKT1*/*PTEN*-altered tumors^32^.

Subprotocol Z1A tested the MEK inhibitor binimetinib in NRAS mutant tumors, which was overall ineffective with an ORR of 2%^24^ (**Figure 1G**). Patients with cholangiocarcinoma (denoted biliary; n=6) exhibited a statistically significant difference in tumor shrinkage compared to all patients (−9.8% versus trial average +6.6%; *P* = 0.047); although the absolute magnitude of tumor shrinkage in this group remains poor.

Subprotocol B tested the HER2 family inhibitor afatinib in tumors with HER2 activating mutations, and had an ORR of 2.7%^27^ (**Figure 1H**). Patients with ER+ breast cancer (n=10) demonstrated a nominally greater tumor volume change versus all patients (−3.3% versus trial average +6.4%; *P* = 0.091; this passes the cut-off in the Benjamini-Hochberg procedure). However, both the magnitude of the difference and the absolute magnitude of tumor shrinkage was small.

No statistically significant differences between tumor types were observed in subprotocols Z1D, Q, I, and Z1F, which tested (respectively) nivolumab in mismatch-repair deficient non-colorectal cancers (ORR 36%)^20^, ado-trastuzumab emtansine in *HER2*-amplified tumors excluding breast and gastric adenocarcinoma (ORR 6%)^19^, taselisib in *PIK3CA*-mutated tumors (ORR 0%)^25^, and copanlisib in *PIK3CA*-mutated tumors (ORR 16%)^26^ (**Figure 1D, J, E, F**). In the case of subprotocol Z1D, the absence of detectable differences among tumor types in 42 patients is preliminary support of tumor-agnostic activity by nivolumab (**Figure 1D**), consistent with the tumor-agnostic activity of pembrolizumab in mismatch-repair deficient tumors^8^, which was previously confirmed by permutation testing^18^.

## Discussion

Our secondary analysis of the NCI-MATCH basket trial uncovered evidence in six of ten subprotocols that a specific tumor type was significantly more sensitive to targeted therapy than other types. Multi-histology trials have the potential to identify tumor-agnostic efficacy of precision cancer medicines, as was achieved by subprotocol H of NCI-MATCH (dabrafenib plus trametinib for *BRAFV600E* tumors)^14^. However, most existing cancer therapies are effective for specific tumor types, and when this is true of a therapy in a basket trial, a low response rate among diverse tumor types is expected. In such cases, secondary analysis by permutation testing can identify drug sensitive tumor types. Indeed, many subprotocols in NCI-MATCH had low response rates, yet most demonstrated substantial efficacy in select tumor types, with average tumor shrinkage of -22% to -60% in five such groups.

With seven significant signals from ten subprotocols, these findings from NCI-MATCH comprise a signal discovery rate that compares favorably to phase 2 trials in oncology in general^33^. Though the generally low response rates in NCI-MATCH were met with some pessimism^13^, our findings suggest that even when a search for tumor-agnostic efficacy is negative, tumor-specific efficacy can often be found instead. The prevalence of tumor-specific efficacy suggests that combining histology and genetics may be the most informative way to guide precision oncology. We therefore anticipate that basket trials will continue to be essential to identify which tumor types are responsive to molecularly targeted therapies.

These findings highlight the utility of permutation testing to enhance the insights gained from basket trials. Binomial tests of response rate discard quantitative data by dichotomizing patient responses. Conversely, permutation testing analyzes magnitudes of response which provides greater statistical power^18^. Given that multi-histology basket trials often have small samples per tumor type, permutation testing maximizes the insights gained from such trials, especially in settings with low patient accrual such as pediatric and rare diseases^34,35^.

Analyzing subgroups in basket trials has limitations. These analyses are hypothesis-generating, not confirmatory, and its findings are associated with a non-zero false discovery rate. This method aims to detect significant signals from small cohorts (≤20 patients per tumor type) to inform decision-making about future trials. However, testing for differences among tumor types is a separate question from testing absolute efficacy, and such differences need to be interpreted in the context of net efficacy. For example, in trials with a high overall response rate, some tumors may be more responsive than others, even while efficacy across all tumors could justify tumor-agnostic approval. Permutation testing is most helpful in the case of low to intermediate response rates to identify specific tumor types with greater sensitivity. In the context of overall low drug activity, greater sensitivity of one tumor type may not be sufficient to warrant further study. For example, the two weakest signals detected in our study were average tumor volume changes of -10% in *NRAS*-mutant cholangiocarcinoma treated with binimetinib, and -3% in HER2-mutant ER+ breast cancers treated with afatinib. Across all subprotocols, few statistically significant differences in PFS were observed, consistent with our prior study^18^. We anticipate that larger cohorts are required to identify differences in PFS.

In conclusion, permutation testing identified many instances where specific tumor types were significantly more treatment-sensitive than others in the NCI-MATCH basket trial. This finding challenges the pessimism about the outcomes of multi-histology basket trials and supports the use of basket trials to identify tumor-specific uses of molecular-targeted therapies. By uncovering promising therapeutic signals in the majority of analyzed therapies, our study suggests that existing targeted therapies may have activity across a range of specific tumor types, which highlights the potential for precision oncology to benefit patients using both histological and genetic information.

## Data Availability

All data analyzed in the present work was previously published, as cited.

## Acknowledgements

D.P. is supported by NIGMS grant T32-GM007753 and F30-CA260780.

