## Supplementary Table 1 for "Tumor-specific activity of precision medicines in the NCI-MATCH trial"

### Supplementary Table 1. NCI-MATCH subprotocols with published results

| # | Journal | Subprotocol | Title | Objective Response Rate (ORR) |
|---|---|---|---|---|
| **1** | Annals of Oncology | Q | Ado-trastuzumab emtansine (T-DM1) in patients with HER2 amplified tumors excluding breast and gastric/gastro-esophageal junction (GEJ) adenocarcinomas: Results from the NCI-MATCH Trial (EAY131) Subprotocol Q<br>Jhaveri KL. *Ann Oncol*. 2019; 30:1821 | 5.6% |
| **2** | Journal of Clinical Oncology | Z1D | Nivolumab Is Effective in Mismatch Repair–Deficient Noncolorectal Cancers: Results From Arm Z1D—A Subprotocol of the NCI-MATCH (EAY131) Study<br>Azad NS. *J Clin Oncol*. 2020; 38:214 | 36% |
| **3** | Clinical Cancer Research | R | Trametinib activity in patients with solid tumors and lymphomas harboring BRAF non-V600 mutations or fusions: Results from NCI-MATCH (EAY131)<br>Johnson DB. *Clin Cancer Res*. 2020; 26:1812 | 3% |
| **4** | Journal of Clinical Oncology | W | Phase II Study of AZD4547 in Patients With Tumors Harboring Aberrations in the FGFR Pathway: Results From the NCI-MATCH Trial (EAY131) Subprotocol W<br>Chae YK. *J Clin Oncol*. 2020; 38:2407 | 8% |
| **5** | Journal of Clinical Oncology | H | Dabrafenib and Trametinib in Patients With Tumors With BRAFV600E Mutations: Results of the NCI-MATCH Trial Subprotocol H<br>Salama AKS. *J Clin Oncol*. 2020; 38:3895 | 37.5% |
| **6** | JAMA Oncology | Y | Effect of Capivasertib in Patients with an AKT1 E17K-mutated Tumor: NCI-MATCH Subprotocol EAY131-Y Nonrandomized Trial<br>Kalinsky KM. *JAMA Oncol*. 2021; 7:271 | 28.6% |
| **7** | Clinical Cancer Research | Z1A | Differential Outcomes in Codon 12/13 and Codon 61 NRAS-Mutated Cancers in the Phase 2 NCI-MATCH Trial of Binimetinib in Patients with NRAS-Mutated Tumors<br>Cleary JM. *Clin Cancer Res*. 2021; 27:2996 | 2.1% |
| **8** | Journal of Clinical Oncology | I | Phase II Study of Taselisib in PIK3CA Mutated Solid Tumors Other Than Breast and Squamous Lung Cancer: Results From the NCI-MATCH ECOG-ACRIN Trial (EAY131) Subprotocol I<br>Krop IE. *JCO Precis Oncol*. 2022;6:e2100424 | 0% |
| **9** | Journal of Clinical Oncology | Z1F | Phase II Study of Copanlisib in Patients With Tumors With PIK3CA Mutations: Results From the NCI-MATCH ECOG-ACRIN Trial (EAY131) Subprotocol Z1F<br>Damodaran S. *J Clin Oncol*. 2022; 40:1552 | 16% |
| **10** | NPJ Precision Oncology | F, G | Crizotinib in patients with tumors harboring ALK or ROS1 rearrangements in the NCI-MATCH trial<br>Mansfield AS. *NPJ Precis Oncol.* 2022; 6:13 | 50%, 25% |
| **11** | Journal of Clinical Oncology | B | Phase II Study of Afatinib in Patients With Tumors With Human Epidermal Growth Factor Receptor 2–Activating Mutations: Results From the National Cancer Institute–Molecular Analysis for Therapy Choice ECOG-ACRIN Trial (EAY131) Subprotocol EAY131-B<br>Bedard PL. *JCO Precis Oncol*. 2022; 6:e2200165 | 2.7% |
